# Multivariable risk modelling and survival analysis with machine learning in SARS-CoV-2 infection

**DOI:** 10.1101/2023.06.22.23291773

**Authors:** Andrea Ciarmiello, Francesca Tutino, Elisabetta Giovannini, Amalia Milano, Matteo Barattini, Nikola Yosifov, Debora Calvi, Maurizio Setti, Massimiliano Sivori, Cinzia Sani, Andrea Bastreri, Raffaele Staffiere, Teseo Stefanini, Stefania Artioli, Giampiero Giovacchini

## Abstract

**Aim:** We evaluated the performance of a machine learning model based on demographic variables, blood tests, pre-existing comorbidities, and CT-based radiomic features to predict critical outcome in patients with acute respiratory syndrome coronavirus 2 (SARS-CoV-2).

**Methods:** We retrospectively enrolled 694 SARS-CoV-2 positive patients. Clinical and demographic data were extracted from clinical records. Radiomic data were extracted from CT. Patients were randomized to the training (80%, n=556) or test (20%, n=138) dataset. The training set was used to define the association between severity of disease and comorbidities, laboratory tests, demographic and CT-based radiomic variables, and to implement a risk prediction model. Models were evaluated using the C statistic and Brier scores. The test set was used for external validation.

**Results:** Patients who died (n=157) were predominantly male (66%) over the age of 50 with (median [range] C-reactive protein (CRP)=5 [1, 37] mg/dL, lactate dehydrogenase (LDH)=494 [141, 3,631] U/I and D-dimer=6.006 [168, 152.015] ng/ml). Surviving patients (n=537) had (median [range]) CRP=3 [0, 27] mg/dL, LDH=484 [78, 3.745] U/I, and D-dimer=1.133 [96, 55.660]ng/ml. The strongest risk factors were D-dimer, age, and cardiovascular disease.

The model implemented using the variables identified by the LASSO Cox regression analysis classified 152 of the 157 (97%) non-survivors as high risk individuals (Odd ratio=54.2 [21.9, 134.4]). Median survival in this group (14 [12, 19] days) was not different from that observed in non-survivors (12 [10, 14] days).

**Conclusions:** A machine learning model based on combined data available on the first days of hospitalization (demographics, CT-radiomics, comorbidities, and blood biomarkers), can identify SARS-CoV-2 patients at risk of serious illness and death.

## Introduction

Severe acute respiratory syndrome coronavirus 2 (SARS-CoV-2) disease has had a significant global health and economic impact and continues to be a major concern as new variants are identified and the number of infected people continues to be high [1–3].

The clinical disease phenotype is extremely heterogeneous since the infection can proceed asymptomatically or evolve through forms of different intensity up to severe disease associated in this latter case with a low survival rate [4].

The variability of clinical manifestations makes prediction of outcome particularly difficult and this may be a major issue when the volume of patients is high and resources are limited as it occurs during a pandemic. Therefore, the identification of major risk factors and the implementation of an outcome prediction model could support treatment planning and optimal resource allocation.

To date, several studies have reported the association between the mortality rate and subject’s age, pre-existing comorbidities, some blood biomarkers, and the degree of lung involvement mainly based on CT scans [4–6].

The data published so far have highlighted a greater frailty of adults who would seem to have a higher rate of severe disease and mortality than young patients did [7].

There is broad agreement among the authors that comorbidities are present in approximately half of patients with SARS-CoV-2. According to the paper by Richardson et al. coronary heart disease, hypertension, diabetes and chronic obstructive lung disease are significantly associated with increased mortality [4, 8].

Several blood biomarkers have been associated with SARS-CoV-2. High D-dimer levels have been reported as predictors of mortality in hospitalized patients [9].

Similarly some blood biomarkers of inflammation such as C-reactive protein (CRP) and cell damage such as lactate dehydrogenase (LDH) would appear to be significantly increased in the most severe forms of the disease [10].

The degree of lung involvement as mainly assessed by computed tomography (CT) is a potential predictor of outcome [6, 11–15]. Data of potential clinical interest contained in the medical images can be read by expert radiologists or extracted by means of dedicated software, This approach, is known as radiomics. Recently, several studies have proposed radiomics and deep learning methods to recognize normal lung parenchyma from that affected by SARS-CoV-2 pneumonia [16] or to predict patient diagnosis [17] and outcome [18]. These approaches provide high diagnostic performance, as evidenced by the area under the receiver operating characteristic curve (AUC>=89%) [18, 19].

The current study is aimed at implementing and validating a mortality risk prediction model for SARS-CoV-2 based on demographic data, blood biomarkers, baseline comorbidities and radiomic CT data using machine learning methods.

## Matherial and Methods

### Population

This retrospective study was based on clinical records from patients admitted to hospital services through to the emergency department with fever, sore throat, dry cough, diarrhea, loss of taste or smell chest pain, and/or shortness or breathing difficulty between March 1, 2020 to December 31, 2020. CT images were retrieved from the hospital picture archiving and communications systems (PACs). The regional review committee granted ethical approval (CER Liguria: 251/2020). The written informed consent for this study was waived. Data were de-identified to avoid any potential breach of patient privacy. Inclusion criteria included: i) positive RT-PCR assay for COVID-19; ii) at least one non-contrast chest CT. For patients with multiple RT-PCR test or CT scan the test closest to the time of initial presentation to the emergency department was used. Exclusion criteria included: i) incomplete clinical records ii) evidence of artifacts affecting image quality. Since they did not meet the inclusion criteria 95 of the 789 patients (12%) initially recruited from the institutional database were excluded. Therefore, the study cohort consisted of 694 subjects with RT-PCR confirmed diagnosis of COVID-19 pneumonia encompassing 447 males and 247 females.

Overall survival (OS) was defined as the time from first hospital presentation to date of death or censoring. Patients who were alive were censored at last follow-up to 31st December 2020. The hospital records were used to determine the status of patients.

### CT imaging acquisition and interpretation

All patients underwent non-enhanced chest CT imaging. Images were acquired in supine position on Aquilion; (Toshiba Medical Systems, Tokyo, Japan) and Optima CT660 (GE Healthcare, Milwaukee, WI) multi-detector CT scanner. Following acquisition parameter were used for all scans: tube voltage: 120 kVp, automatic tube current: (120-440) mAs, thickness: (5-7) mm, slice interval: 5 mm, rotation speed: 0.5-1.0 s, helical pitch 1.0875:1 or 1.375:1. Images were reconstructed at 512×512 pixels with a section width of 0.625 mm.

All CT images were reviewed in the imaging laboratory of S. Andrea Hospital by two board-certified radiologists specifically skilled in thoracic imaging. CT images were classified according to the criteria proposed by the Radiological Society of North America (RSNA) [20 302]. Subjects were grouped into two classes defined as typical and atypical findings which included typical / indeterminate and atypical / negative CT patterns as defined by Simpson and colleagues [20 302].

### Image analysis and texture features extraction

Lung images were segmented using the 3D slicer software v4.11 [21]. All segmented images were reviewed by two certified radiologists to rule out those with segmentation errors. Radiomics package was used to extract image feature (https://github.com/mvallieres/radiomics/). CT images were evaluated with the first and second-level order features describing the pattern of spatial distribution of voxels intensity. Texture features consisted of 3 histogram-based, 9 Gray-Level Co-occurrence Matrix (GLCM), 13 Gray-Level Run-Length Matrix (GLRLM), 13 Gray-Level Size Zone Matrix (GLSZM), and 5 Neighborhood Gray-Tone Difference Matrix (NGTDM) features [22, 23]. Therefore the set of radiomic predictors used for this study included 43 features extracted from each CT lung image.

### Feature selection and classification

Least absolute shrinkage and selection operator (LASSO) Cox regression analysis [24] was used to build the model for predicting overall patient survival with the clinical and radiomic features [25]. Regularized Cox models regression was performed with cv.glmnet under R 4.1.3 (http://www.r-project.org) glmnet package [26]. Lasso regularization parameters were chosen by means of the penalty term (L1-norm) tuned with the constant lambda (λ). Tuning parameters were selected using cross-validation to find the λ value able to minimize the mean squared error of the predictions. The cv.glmnet function provides the cross-validated mean C-index and C-index standard error estimate. The function also reports the minimum mean cross-validated error (lambda.min) and the value of lambda providing the most regularized model with a cross-validated error within 1 standard error of the minimum. [25].

### Model design

Fifty-seven predictors were included in Lasso’s initial selection. They included 2 demographic (age and gender), 3 laboratory tests (C-Reactive Protein, Lactate Dehydrogenase and D-dimer), 9 comorbidities (Cancer, blood cancer, diabetes, obesity, hematologc disease, cardiovascular disease, cerebrovascluar disease, chronic obstructive pulunary disease) and 43 radiomic features. The predictive model was implemented with the demographic, metabolic and radiomic characteristics that survived the Lasso analysis with the Cox multiple regression method.

To evaluate the model’s performance on new data not used for training, the cohort was randomly divided to include 80% of the sample in the training set and 20% in the validation set. The proportions for groups splitting has been reported to depend on the sample size and the percentage of complete data. The split ratio used for training and test dataset division has proven accurate in developing predictive models when sample sizes are => 100 and the percentage of cases with a complete data set used for the model estimate is greater than 85% [27].

### Model validation and calibration

The predictive ability of Cox fitted model was evaluted with calibrated and validated functions available under rms package. This platform provide a robust approach including cross validation, bootstrap, randomization and resampling [28]. Calibration method was used to evaluate the performance of the prediction model by comparing the predicted to the observed probabilities. To reduce overfitting and quantify optimism, the model was internally validated by computing an optimism-corrected C-statistic after 1000 bootstrapped resampling. External validation was also performed using a test dataset splitted from the study sample and not used for model training. Model calibration and validation was based on C-index and Brier score metric.

After validation patient’s individual risk score was calculated using the ggrisk package. Subjects were grouped into high- and low-risk groups based on the median risk score. The ability of the risk score to assess the probability of survival was assessed in the whole sample using Kaplan-Meier analysis with ggsurvplot function and log-rank test.

### Statistics

R software (version 4.1.3, http://www.r-project.org) was used to data analysis and graphics. Continuous data were tested using independent t-tests, with degrees of freedom adjusted for inequality of variance where appropriate. Possible association of predictors with patient outcome was assessed with Wald’s test [29, 30]. LASSO logistic regression analysis was conducted using the glmnet package in R. The survival curves were generated using the Kaplan-Meier method implemented in the ggsurvplot function. The pROC and survival-ROC packages were applied to analyze ROC curves. Validation plots were produced by the root mean squares (RMS) package. The chi-square analysis was used for categorical variables. Sensitivity (SS) and specificity (SP) odd ratio (OR) and their 95% confidence intervals (CIs), were calculated to estimate how strongly the model predicted diagnosis was associated with clinical outcome. Two-tailed P values of less than 0.05 were considered statistically significant.

## Results

A total of 694 patients were recruited and randomized to include 80% (n = 556) in training and 20% (n = 138) in test datasets. Patient’s characteristics were summarized in Table 1. The median age was 64 years (range: 20-107). The study sample consisted predominantly of males (64%). The majority of patients were resident in northeastern Italy. Median hospital stay was 11 days (range: 3-86). Patients had median CRP of 3 mg/dL (range: 0.11-37) and median LDH of 48 U/I (range: 78-3745). Moreover, patients had a median d-dimer of 1133 ng/mL (range: 96-152015). Deceased patients were predominantly male (66%) older than 50 years. Compared with survivors, deceased patients showed differences in laboratory findings (Table 1). As expected, also in the current study sample D-dimer, CRP and LDH were significantly increased in non-survivors compared with survivors (Table 1). Visual assessment of CT images according to RSNA guidelines [20], identified 111 of 157 non-survivors and 299 of 537 survivors with typical findings.

**Table 1.** Clinical characteristics of Sars-CoV-2 population in alive and deceased subjects.

| Variable | Overall, N = 694<sup>1</sup> | Observed outcome |  | Statistic | p-value<sup>2</sup> |
| --- | --- | --- | --- | --- | --- |
|  |  | Alive, N = 537 | Deceased, N = 157 |  |  |
| **Age** |  |  |  | 213 | **<0.001** |
| Median (Range) | 64 (20 - 107) | 59 (20 - 94) | 80 (51 - 107) |  |  |
| **Gender, n (%)** |  |  |  | 0.07 | 0.8 |
| Female | 247 (36%) | 193 (36%) | 54 (34%) |  |  |
| Male | 447 (64%) | 344 (64%) | 103 (66%) |  |  |
| **Hospital stay** |  |  |  | 10 | **0.001** |
| Median (Range) | 11 (3 - 86) | 10 (3 - 86) | 13 (3 - 62) |  |  |
| **CT-findings, n (%)** |  |  |  | 11 | **0.001** |
| Negative/Atypical | 284 (41%) | 238 (44%) | 46 (29%) |  |  |
| Typical | 410 (59%) | 299 (56%) | 111 (71%) |  |  |
| **C-reactive protein** |  |  |  | 19 | **<0.001** |
| Median (Range) | 3 (0 - 37) | 3 (0 - 27) | 5 (1 - 37) |  |  |
| **Lactate dehydrogenase** |  |  |  | 97 | **<0.001** |
| Median (Range) | 486 (78 - 3,745) | 484 (78 - 3,745) | 494 (141 - 3,631) |  |  |
| **D-dimer** |  |  |  | 68 | **<0.001** |
| Median (Range) | 1,133 (96 - 152,015) | 1,133 (96 - 55,660) | 6,006 (168 - 152,015) |  |  |

<sup>1</sup>n (%)
<sup>2</sup>One-way ANOVA; Pearson's Chi-squared test

Table 2 shows the impact of pre-existing comorbidities on mortality in patients with SARS-CoV-2. In particular, cardiovascular and cerebrovascular diseases, cancer, haematological diseases and chronic obstructive pulmonary disease significantly increase the probability of death in the study sample.

**Table 2.** Diseases associated with a high risk of mortality in SARS-CoV-2 infection.

| Comorbidity | Alive, N = 537 | Deceased, N = 157 | Odds Ratio<sup>1,2</sup> | p-value<sup>1</sup> |
| --- | --- | --- | --- | --- |
| **Cardiovascular Disease, n (%)** | 90 (17%) | 74 (47%) | 4.42 (2.95 to 6.63) | **<0.001** |
| **Cancer, n (%)** | 2 (0%) | 9 (6%) | 16.2 (3.30 to 155) | **<0.001** |
| **Cerebrovascular Disease, n (%)** | 9 (2%) | 14 (9%) | 5.72 (2.25 to 15.3) | **<0.001** |
| **Hematologic Disease, n (%)** | 14 (3%) | 13 (8%) | 3.36 (1.42 to 7.91) | **0.003** |
| **Chronic obstructive pulmonary disease, n (%)** | 24 (4%) | 16 (10%) | 2.42 (1.17 to 4.90) | **0.011** |
| **Blood Cancer, n (%)** | 11 (2%) | 1 (1%) | 0.31 (0.01 to 2.14) | 0.32 |
| **Hypertension, n (%)** | 66 (12%) | 16 (10%) | 0.81 (0.42 to 1.47) | 0.57 |
| **Type 2 Diabetes, n (%)** | 77 (14%) | 20 (13%) | 0.87 (0.49 to 1.50) | 0.70 |
| **Obesity, n (%)** | 11 (2%) | 3 (2%) | 0.93 (0.16 to 3.59) | >0.99 |

<sup>1</sup>Fisher's Exact Test for Count Data
<sup>2</sup>CI = Confidence Interval

Figure 1 shows the relevance of each predictor based on the Wald test obtained from multivariable logistic regression used for modeling patient’s mortality. Predictors are sorted by decreasing importance and only those with a significance <= 0.05 are shown. The most important predictor was the D-dimer which resulted the most significant among the laboratory tests and the demographic variables used to define the prediction model. Moreover, important outcome predictors were also found among some textural features belonging to the GLOBAL, GLCM, GLSZM, GLRLM and NGDTM families. Among the comorbidities, cardiovascular disease appears to have a significant impact on survival, ranking among the most significant predictors of mortality.

**Figure 1.**
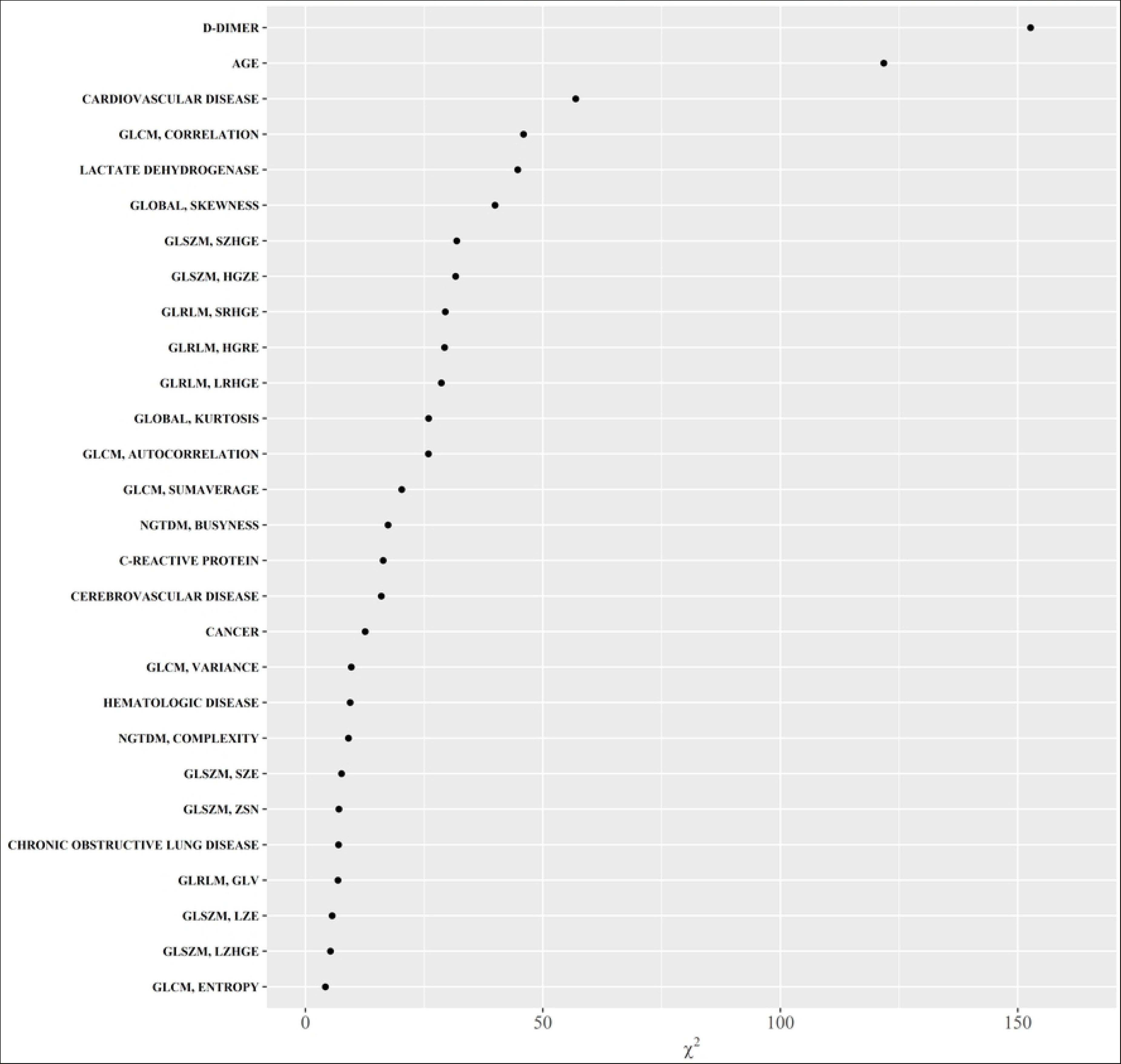
Predictors importance. Relevance of predictors based on the Wald test obtained from multivariable logistic regression. Predictors are shown by decreasing statistical importance, and only those with a significance ≤ 0.05 are shown. The D-dimer was the most important predictor. Important predictors were several textural features belonging to the GLOBAL, GLCM, GLSZM, GLRLM and NGDTM families. Among comorbidities, cardiovascular disease was the strongest risk factor

Based on the results of the LASSO regression a mixed model to predict survival in hospitalized SARS-COV-2 patients was implemented. Prediction model optimization was based on C-index using 10-fold cross validation. The parameter tuning producing a C-index within 1 standard error was 0.053 corresponding to a C-index of 0.87 (standard error 0.013) (Figure 2A, B). Twelve out of 57 variables with non-zero coefficients survived the tuning parameters giving the C-index within 1 standard error of the maximum (Figure 2C). Selected variables included age, d-dimer, LDH, 3 groups of comorbidities and 6 radiomic variables (Figure 2C).

**Figure 2.**
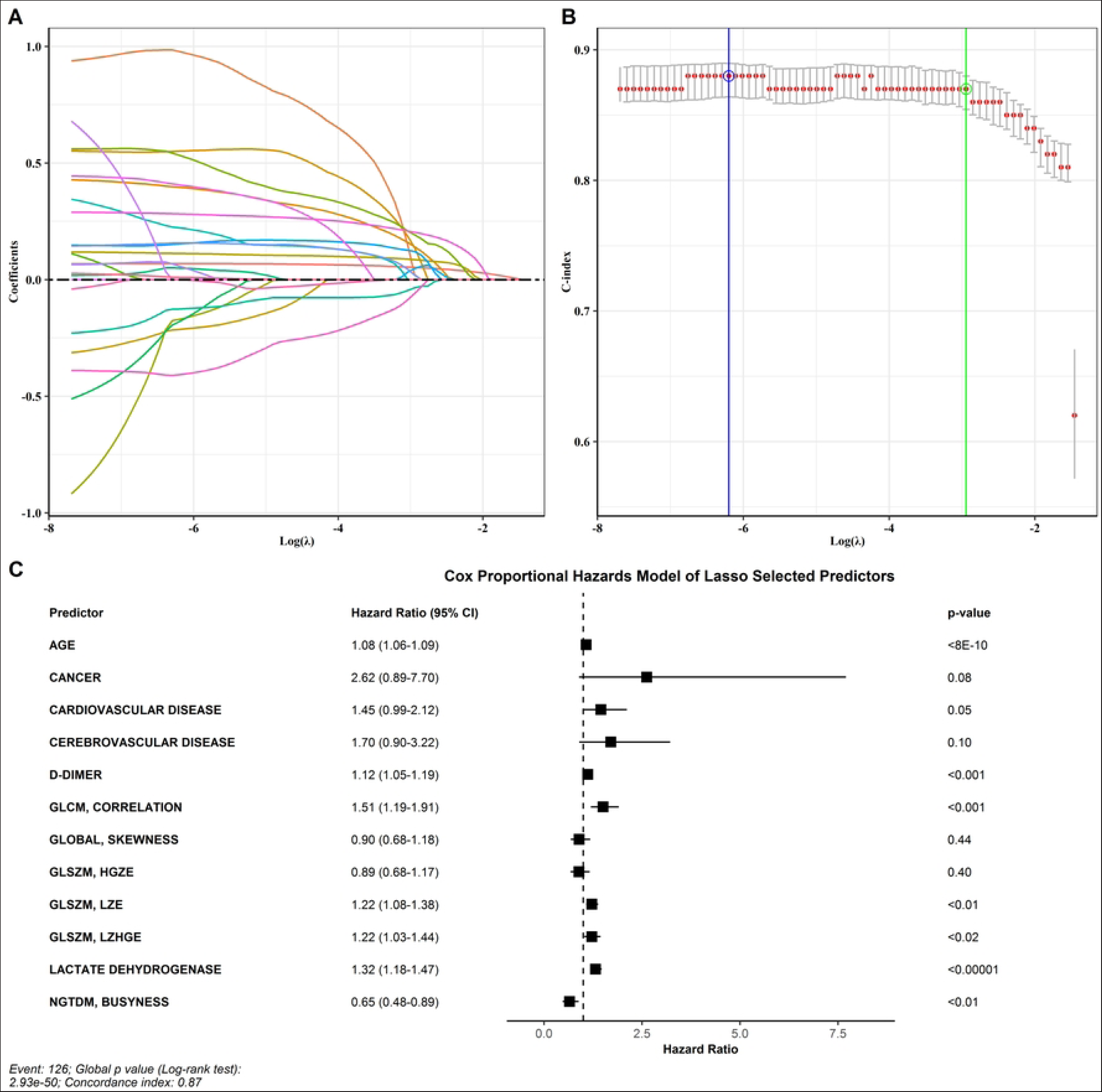
Predictors of outcome. Parameter tuning (A). C index (B). Variables that survived at the LASSO regression, including age, D-dimer, LDH, 3 comorbidities and 6 radiomic variables (C)

Internal validation showed high agreement between the predicted and observed survival curves (Figure 3 left panel). The unadjusted and bias-adjusted curves were similar and aligned with the dashed curve representing the best possible relationship between observed and predicted outcome as estimated by the mean absolute error (MAE) of 0.03. Furthermore, the C statistic and Brier score used as a measure of prediction accuracy were also 0.893 and 0.068, respectively, confirming significant agreement between the estimates. A sample of 138 SARS-COV-2 patients was randomly drawn from the study population and used as a test set for external validation. The mean absolute error estimated between the predicted and observed curves in the test set was 0.05 (Figure 3 right panel). The C-index and Brier score on this dataset were 0.886 and 0.050, respectively.

**Figure 3.**
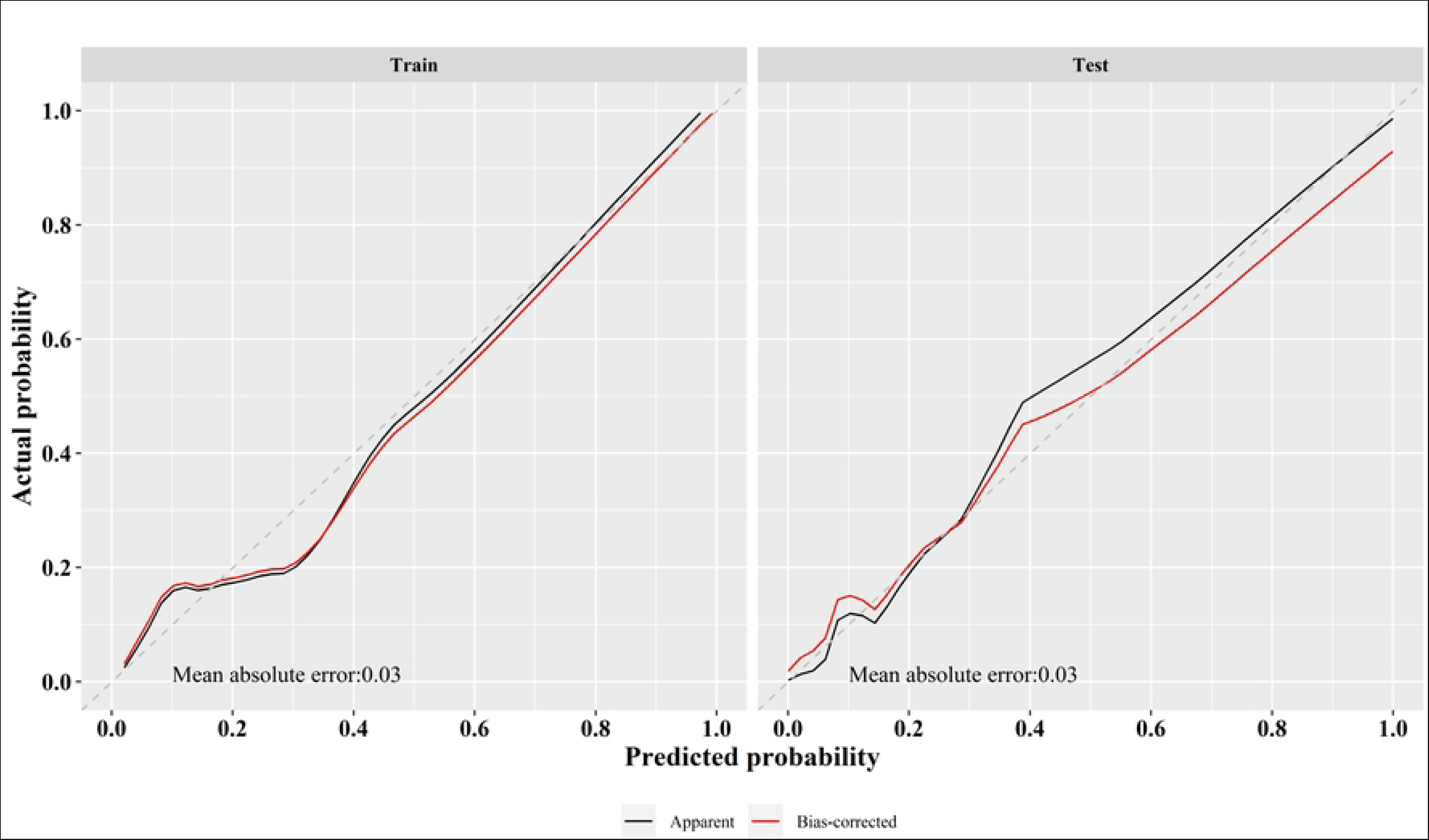
Calibration curves. Internal validation (left panel) shows high agreement between the predicted and observed survival curves. The unadjusted and bias-adjusted curves were similar to the dashed curve representing the best possible relationship between observed and predicted outcome as estimated by the mean absolute error (MAE) of 0.03. In the test set for external validation (right panel), the MAE between the predicted and observed curves was 0.03

Using the Cox regression algorithm on the predictors selected by the operator Lasso, the individual risk score of the patients included in the training dataset was estimated. The median risk score of 0.18 was used as a cutoff point, to split patients into high- and low-risk groups. KM survival analysis was subsequently performed to evaluate the predictor accuracy and build a risk model for the survival rate of SARS-COV-2 patients in both the train and test datasets.

In the training data set, median survival was 12 days (95% CI; 10-14). Using the mixed model for risk prediction in the training dataset, the survival time of SARS-COV-2 patients with a 50% survival rate in the high-risk group was significantly different compared to the low-risk subjects (Log-rank; p<0.001), with a median of 15 days (95%CI; 12-20) (Figure 4A). By contrast, the low-risk group did not achieve the 50% survival rate.

**Figure 4.**
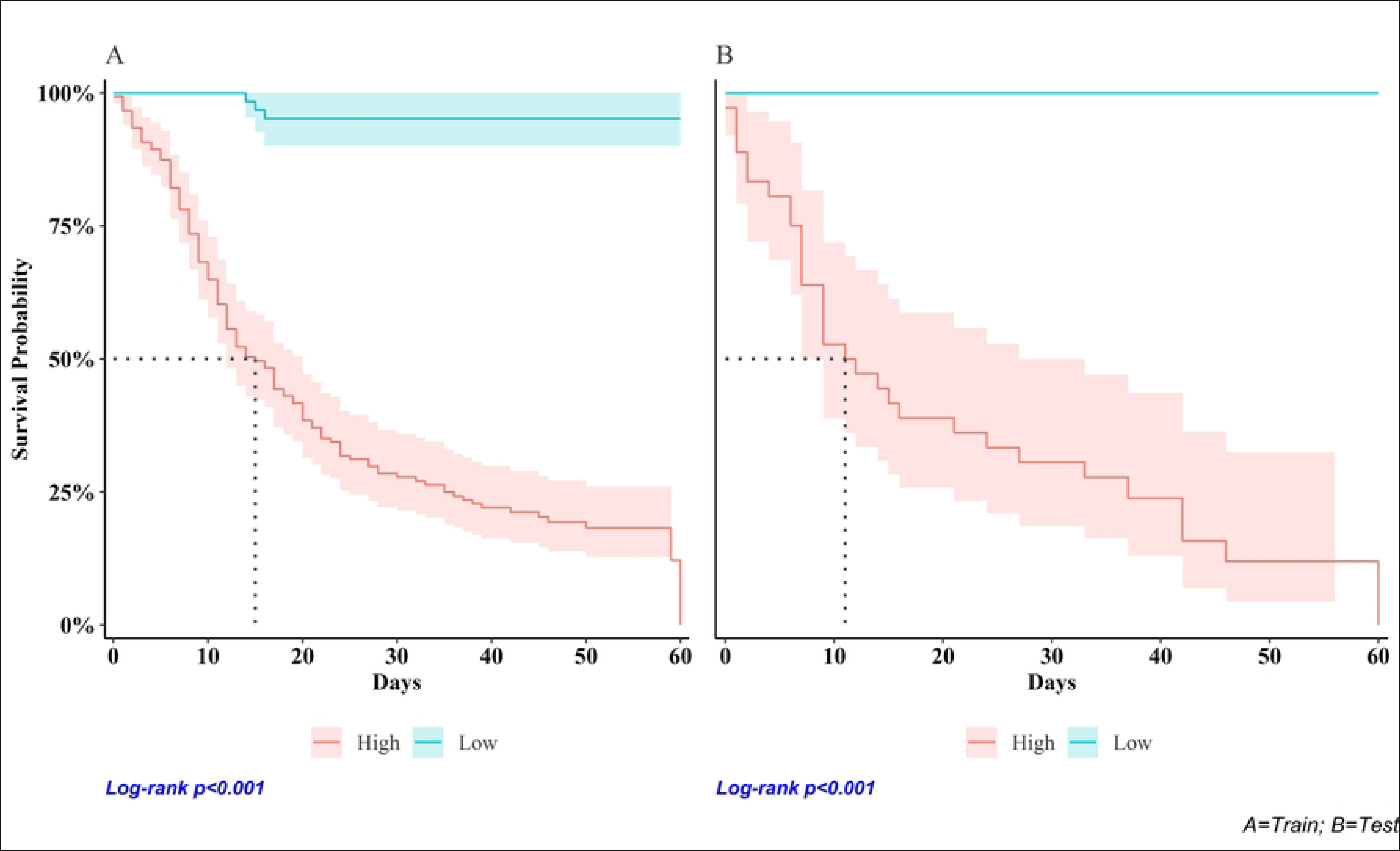
Survival curves. Training data set (A): the survival time of SARS-CoV-2 patients in the high-risk group was significantly different compared to the low-risk subjects, with a median of 15 days (95%CI; 12-20). The low-risk group did not achieve the 50% survival rate. Test dataset (B): the median survival of the high risk group was 12 days (95% CI; 7-27) and low-risk patients did not reach the median survival of 50%

The median survival observed in the test dataset was 9 days, (7-16). In the test dataset, the observed trend was similar to that estimated in the training group with a median survival of 12 days (95% CI; 7-27). As in the training sample, also in the test dataset, low-risk patients did not reach the median survival of 50% (Log-rank p<0.001) (Figure 4B).

Table 3 shows the comparison between the observed outcome and the expected risk. The prediction model identifies 97% of true positives among subjects at risk of death while 64% of true negatives were classified as having a low risk of event. The risk of mortality was found to be significantly higher (odds ratio = 54.2 (95%CI; 21.9-134.4), p<0.0001) among the high-risk group compared to the low-risk group.

**Table 3.** Bivariate analysis of predicted risk on disease outcome.

| Predicted | Observed |  | X<sup>2</sup> | p-value<sup>1</sup> | SS 95%CI<sup>2</sup> | SP 95%CI<sup>3</sup> | OR 95%CI<sup>4</sup> |
| --- | --- | --- | --- | --- | --- | --- | --- |
|  | Deceased, N = 157 | Alive, N = 537 |  |  |  |  |  |
| Mixed, n (%) |  |  | 178 | 1.6e-40 | 97 (94, 100) | 64 (60, 68) | 54.2 (21.9, 134.4) |
| High risk | 152 (97%) | 193 (36%) |  |  |  |  |  |
| Low risk | 5 (3.2%) | 344 (64%) |  |  |  |  |  |

<sup>1</sup>Pearson's Chi-squared test
<sup>2</sup>Sensitivity, Confidence Interval
<sup>3</sup>Specificity, Confidence Interval
<sup>4</sup>Odd Ratio, Confidence Interval

## Discussion

Prediction of disease severity and progression in SARS-CoV-2 patients is relevant as early intervention is significantly associated with reduced mortality [31, 32]. In this study, we developed and validated a risk scoring model based on demographics, laboratory tests, and radiomic features to predict the disease progression and survival of hospitalized patients with SARS-CoV-2.

The model was implemented with 12 out of 57 variables selected using Lasso Cox regression and the C-index metric. The proposed model is highly predictive, identifying 97% of deceased patients as high-risk and 64% of surviving patients as low-risk. In addition, how well the model-predicted risk describes the observed sequence of events is summarized by the estimated C-index of 0.90.

The risk estimation model included age, laboratory tests (D-dimer, LDH) and 7 radiomic features. The variables used to estimate the risk of developing critical illness due to SARS-CoV-2 infection are generally available during the early stages of hospitalization. Risk estimation in this phase could support clinicians in planning the treatment strategy by allocating resources for more aggressive treatments or admission to intensive care units for higher risk cases or by choosing to watch and wait for low risk cases.

Previous studies have reported the impact of age on SARS-CoV-2 mortality. A meta-analysis demonstrated a decisive effect of age on mortality [7]. A 60% higher risk of mortality was reported in subjects aged >80 years [7]. As expected also in our study the age of non-surviving patients was significantly higher than survivors (median 80 years, 95%CI: 51-107 vs. 59yo, 95%CI: 20-94; p<0.001). Age was an important predictor of disease outcome and survived the Lasso regression thus contributing to prediction model implementation.

D-dimer was associated with poor outcome of patients with SARS-CoV-2, presumably due to the increased likelihood of developing pulmonary embolism with D-dimer levels above 2590 μg/ml [33]. According to recently published papers [34, 35], also in our sample, D-dimer was the variable with the strongest association with patient outcome as suggested by measured levels of 6.006 vs 1.133 μg/ml for deceased and survivors, respectively.

Similarly, elevated lactate dehydrogenase (LDH) levels have been associated with worse outcomes in patients with viral infections [36, 37]. Deceased patients in our study had significantly higher LDH levels than survivors and it was selected by Lasso regression for survival prediction model.

Literature reports have documented that chronic comorbidities are associated with increased risk of poor prognosis and fatal outcome associated with SARS-CoV-2 [8]. Similarly, in our model, pre-existing comorbidities, like cardiovascular and cerebrovascular diseases, cancer, hematological diseases and chronic obstructive pulmonary disease, were significant predictors of severity of disease and death. Among comorbidities, cardiovascular disease resulted the strongest predictor of mortality in our study sample with a 4.42 fold higher risk of poor prognosis, in line with findings of meta-analyses [38–41].

CT is the most widespread imaging modalities that play a key role in the diagnosis and assessment of prognosis in patients with SARS-CoV-2 [42]. However, CT findings (such as ground-glass opacities, consolidation) are not specific for SARS-CoV-2, as similar findings can also be found in other diseases, such as seasonal influenza associated with a lower risk of death.

Innovative methods of quantitative image analysis (such as radiomics) can provide an operator independent semi-quantitative approach by describing spatial and temporal information derived from images (CT, MRI, PET/CT). Until now, radiomics has found application in different scenarios of medicine such as oncology and neurodegenerative disease [43, 44, 45]. Lately it has also been used to support the “digital biopsy,” a non-invasive tissue characterization technique.

Previous studies have reported the potential use of CT radiomic features to better characterize pulmonary involvement of patients with SARS-CoV-2. Spatial information measured with radiomic features can be used to support differential diagnosis between covid and non-covid disease [46] as well as in modeling risk of death and predicting survival[47].

In our study, six radiomic features (Global_Skewness, GLCM_Correlation, GLSZM_LZE, GLSZM_HGZE, GLSZM_LZHGE, NGTDM_Busyness) were selected to model a risk profile with significant discriminative capabilities for patient outcome. Indeed, selected variables were significantly associated with patient outcome in multivariate logistic regression (p<0.001). These features contribute to risk modeling by providing quantitative information on lung CT signal intensity and heterogeneity in SARS-CoV-2 patients.

A systematic review of existing prognostic models identified several prognostic models designed to support diagnosis and predict mortality among SARS-CoV-2 hospitalized patients [5]. Most of the studies reported predictive models implemented with CT images and/or clinical variables combined differently depending on the available data.

Only few studies included radiomics, demographics, comorbidities and laboratory tests together as potential predictor candidates. The main disadvantage of these studies is related to the small sample size which exposes the results to a high risk of bias due to inappropriate evaluation of the predictive performance on external dataset and inappropriate missing data handling.

Our study included 694 patients with complete radiomic and clinical datasets. The predictors needed to calculate the risk of developing serious disease are usually available within the first few hours of hospital admission. Based on these variables, the model is able to estimate the risk of mortality, identifying 97% of non-survivors in the study sample. The availability of this information could be useful for optimizing treatment planning according to the estimated risk during patient admission to hospital.

The major limitation of the study lies in the lack of an external validation carried out on a dataset obtained from another hospital. Although the external validation was performed on a test set not used for training, to build a robust model and to obtain reliable performance evaluation it would be advisable to validate the model on data from different sources.

The model is not available as a ready-to-use software package. The study was designed to define and validate a predictive risk model to be subsequently produced as a usable application in clinical practice. To this end, open source widespread statistical software was used. These packages can easily allow to transfer the method into clinical practice.

## Conclusion

A predictive model of mortality was developed in a sample of 694 SARS-CoV-2 patients using demographic, CT-radiomic and laboratory tests. The model was calibrated and validated by randomly splitting the sample into the training and test dataset. The final model was implemented with a combination of 12 variables including age, D-dimer, LDH, preexisting comorbidities as cancer, cardiovascular and cerebrovascular disease and 6 radiomic features. The model was able to correctly identify 97% of non-survivors. Identifying high-risk individuals with predictors usually available within the first few hours of hospital admission could be useful in case of widespread disease for a better allocation of available resources.

## Data Availability

All relevant data are within the manuscript and its Supporting Information files.

## Acknowledgments

The authors would like to thank all study participants, including Dr. Manuele Sicuteri, head of the information and communication technology unit of the S. Andrea hospital, whose support in the data retrieving and organization was fundamental.

## Notes

### Competing Interest Statement

The authors have declared no competing interest.

### Funding Statement

The authors received no specific funding for this work.

### Author Declarations

All procedures performed in studies involving human participants were in accordance with the ethical standards of the 1964 Helsinki Declaration and its later amendments or comparable ethical standards. The Regional Review Committee granted ethical approval (CER Liguria: 251/2020).

